# Total parietal peritonectomy leads to a low incidence of platinum resistance in patients undergoing interval cytoreductive surgery for advanced epithelial serous ovarian cancer- results of a prospective multi-centric study

**DOI:** 10.1101/2020.06.28.20141903

**Authors:** Praveen Kammar, Snita Sinukumar, Loma Parikh, Sakina Shaikh, Nutan Jumale, Mrinal Mallya, Sandeep Sheth, Sanket Mehta, Aditi Bhatt

## Abstract

**Background and Aim:** Though interval cytoreductive surgery (CRS) following neoadjuvant chemotherapy (NACT) is considered non-inferior to primary CRS, the incidence of platinum resistance is high. A total parietal peritonectomy (TPP) can address occult microscopic disease more completely and could reduce the rates of early recurrence. The early results of a prospective study evaluating the role of TPP are presented in this manuscript.

**Methods:** This is a prospective, multi-centric interventional study. A TPP was performed in all patients undergoing interval CRS. A fixed surgical protocol was followed. Grade 3-4 morbidity was recorded. Factors affecting grade 3-4 morbidity, early recurrence and progression-free survival (PFS) were evaluated.

**Results:** From July 2018, 70 patients with serous carcinoma were included. The median surgical PCI was 15 [range 5-37]. A CC-0 resection was obtained in 55(78.5%) patients; CC-1 in 10(14.2%). Grade 3-4 complications were seen in 15(21.4%) patients of which the commonest complication was intraperitoneal fluid collection. Occult disease in the peritoneum was seen in 40%. Early recurrence (platinum resistance) was seen in 5(7.1%). The median PFS was 18 months [range 0-21months]. Patients with a lower PCI and with no grade 3-4 complications had a significantly longer PFS. A pathological PCI>15 was the only independent predictor of a shorter PFS (p=0.001).

**Conclusions:** TPP performed as a part of interval CRS resulted in a very low incidence of platinum resistance. The post-operative morbidity was acceptable. These findings should be confirmed in a larger series and a randomized trial performed to demonstrate demonstrate its benefit over conventional surgery.

## Introduction

Cytoreductive surgery (CRS) and systemic chemotherapy is the standard of care for patients with advanced ovarian cancer. [1] CRS may be performed after few cycles of neoadjuvant chemotherapy (NACT) in some patients who have extensive disease or in whom major morbidity is anticipated. [2] Though the NACT approach is considered non-inferior to performing CRS upfront, the incidence of platinum resistance is higher. [3] One randomized trial showed the benefit of hyperthermic intraperitoneal chemotherapy (HIPEC) in addition to complete CRS, but the benefit in progression free survival (PFS) was only 3 months. [4] Another strategy that has led to an increase in the progression-free survival has been the use of maintenance therapy with antiangiogenic therapy (beavacizumab) and/or PARP inhibitors. [5, 6, 7] However, the benefit in patients undergoing interval CRS and the impact on platinum resistance has not been studied.

Conventionally, surgery that is performed in the interval setting addresses only sites of visible residual disease and the parietal peritoneal resection can be termed as ‘selective parietal peritonectomy’ (SPP).

Several mechanisms of platinum resistance have been described, but one that is often not considered is the inability to address all the sites of residual disease adequately during interval CRS. [8, 9] Visual inspection has reduced accuracy in determining the presence or absence of disease in these patients due to the scarring produced by NACT. [10] The scar tissue is known to harbor chemotherapy resistant stem cells which are a potential source of recurrence. [11] Complete removal of the parietal peritoneum, that is a total parietal peritonectomy (TPP) can address all these issues and has the benefit of removing the parietal peritoneum that is the more common site of residual disease compared to visceral regions (excluding the omentum). Our previous study showed a low morbidity of a TPP in this setting. [12] Other investigators have developed similar strategies to increase the rates of complete cytoreduction. [13] We present the early results (morbidity and incidence of recurrence within 6 months) of our prospective study addressing the role of a TPP in patients with serous epithelial ovarian cancer undergoing interval CRS.

## Methods

This is a prospective interventional, multi-centric study being carried out at three peritoneal surface malignancy centres. It is registered with the clinical trials registry of India (CTRI/2018/08/015350). The study was approved by the Zydus Hospital Ethics committee on 30^th^ June 2018. Subsequently, approval was obtained at the other participating hospitals. Informed consent was obtained from all patients.

### Study protocol

Patients with stage IIIC and IVA serous epithelial ovarian cancer undergoing interval CRS were included. Patients who had resectable disease (CC-0 resection possible), good performance status (ECOG-0/1) and absence of ascites >500ml were taken up for surgery upfront **(Figure 1)** and thus, excluded. The remaining were reassessed after 3 cycle of neoadjuvant chemotherapy (NACT) comprising of standard doublet regimens (platinum compound and taxane). Patients having a clinical and radiological response and in whom a complete resection was possible were taken up for interval CRS. Response evaluation was based on the RECIST 1.1 criteria. [14] Others were reassessed for resectability after the completion of all 6 cycles. Patients who had stage IV-A disease and negative pleural fluid cytology after NACT are taken up for surgery. Surgery was performed 3-4 weeks after completion of the third cycle. Patients with non-serous histologies, those refusing consent, those with complete bowel obstruction and/or a poor performance status (Eastern Co-operative Oncology Group – ECOG>1) were excluded. Patient who had unresectable disease after 6 cycles or those that did not undergo surgery or those receiving second line chemotherapy were excluded. Since the publication of the Dutch randomized controlled trail, HIPEC in the interval setting has been introduced in the NCCN guidelines which are widely followed in India. [4,15] However, it remains an out-of-pocket expenditure for patients and was performed only for patients who could afford the additional cost and were willing for the procedure. HIPEC was performed with cisplatin 75mg/m2 for 90 minutes by the open (2 centres) or closed method (1 centre). Adjuvant chemotherapy was started for all patients within 4-6 weeks of surgery. Maintenance therapy was at the discretion of the treating physician.

**Figure 1.**
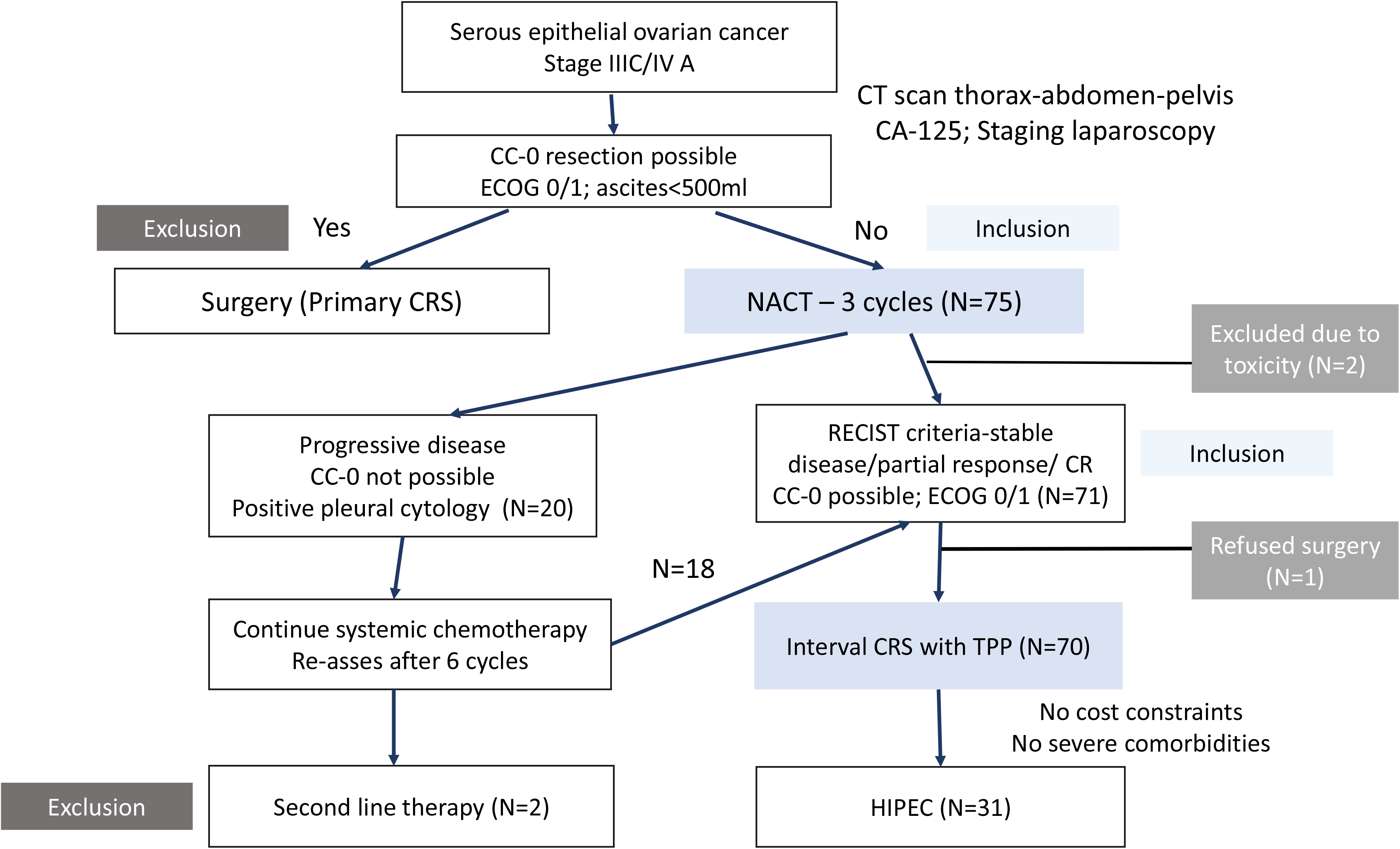
Flow-chart showing inclusion and exclusion of patients in the study at different time points Abbreviations: NACT-neoadjuvant chemotherapy; CRS-cytoreductive surgery; ECOG-Eastern Cooperative oncology group; TPP-total parietal peritonectomy; CR-complete response

In this manuscript, we discuss the early outcomes in patients undergoing interval CRS. Only patients who had completed a minimum of 6 months of follow-up were included.

### Study end points and sample size

The primary end-point of the study is progression-free survival (PFS) and the secondary end-points overall survival (OS), major morbidity and impact of chemotherapy response grade on survival. A median PFS of 15-18 months is obtained with standard surgery (SPP). The expected benefit of TPP in PFS is 30% and to detect this difference with a power of 80% and an alpha of 0.05, 150 patients are required. The sample size was set at 200 for this study.

### Surgical protocol

The goal of surgery was to obtain a complete cytoreduction (CC-0 resection). [16] The detailed protocol is in supplement 1. Briefly, a midline incision from the xiphoid to the pubis was employed irrespective of the disease extent. The disease was quantified using Sugarbaker’s peritoneal cancer index (PCI); this was the surgical PCI (sPCI). [16] TPP comprised of all 5 peritonectomies-pelvic, bilateral antero-parietal, right and left upper quadrant peritonectomies and a total omentectomy. The regions to be resected for each peritonectomy, the anatomical boundaries were described and defined **(supplement 1)**. For mesenteric disease, performance of mesenteric peritonectomy was at the discretion of the surgeon. A minimal of representative biopsy of tumor bearing or normal mesenteric peritoneum (if no disease is present) was performed for each of regions 9-12.

Any viscera that was infiltrated by the tumor was resected to obtain a complete cytoreduction. A bilateral pelvic and retroperitoneal lymphadenectomy (till the level of the renal veins) was performed for all patients in whom lymph nodes are suspicious on imaging or intraoperatively.

### Evaluation of morbidity

The 90-day morbidity and mortality were recorded. The common toxicology criteria for adverse events (CTCAE) version 4.3 classification was used to record the morbidity. [17] Grades 3 and 4 are considered major morbidity.

### Pathological evaluation

Pathological evaluation was performed according to a predefined protocol described elsewhere for the peritoneal disease and primary tumour and involved calculation of the pathological PCI (pPCI). [18, 19, 20] Additional sections were taken from the surrounding normal peritoneum in addition to the tumor nodules at a distance of 5mm or more from the edge of the tumor nodules. The chemotherapy response score of Bohm was used to report the pathological response to systemic chemotherapy. [21]

### Follow-up

Routine 3-monthly follow-up was performed for all patients with CA-125 levels and imaging studies as deemed suitable for the first two years and 6-monthly thereafter. Patients who had normalization of CA-125 following the last cycle of chemotherapy with no other clinical evidence of disease were considered to have had a complete response following first line therapy. Recurrence was diagnosed according to the Gynecologic Cancer InterGroup (GCIC) criteria. [22]

### Statistical methods

Categorical data are described as number (%) and continuous data are expressed as the median and range. Categorical data were compared with the chi-square test. For comparison of median values, non-parametric independent sample t test and for means, the Mann Whitney U test was used. Cox proportional hazard regression was used to describe the association between individual risk factors and PFS and overall survival (OS) both, in terms of hazard ratio and its 95% confidence interval (CI). Multivariate Cox regression was used to assess the impact of risk factors on survival. A p-value of <0.05 was considered statistically significant. Survival was calculated from the date of the last cycle of chemotherapy. Early recurrence or platinum resistance was defined as recurrence within 6 months of completion of the last cycle of chemotherapy.

## Results

From July 2018 onwards 70 patients were included. The median age was 56 years [range 33-70 years]. Further details are in **Table 1**. The median sPCI was 15 [range 5-37]. A CC-0 resection was obtained in 55(78.5%) patients, CC-1 in 10(14.2%) and CC-2 in 5(7.1%). The median pPCI was 9 [range 0-26]. A pathological complete response was seen only in 3(4.2%) patients and a near complete response in 11(15.7%). Twenty-eight (40.0%) had disease in ‘normal appearing’ peritoneum and 31(43.0%) in the peritoneum around tumor nodules. The region wise distribution of occult disease is shown in **Figure 2**. All patients started chemotherapy within 40 days of surgery. The median time was 36 days [range 29-40 days]. None of the patients received any kind of maintenance therapy.

**Table 1.**
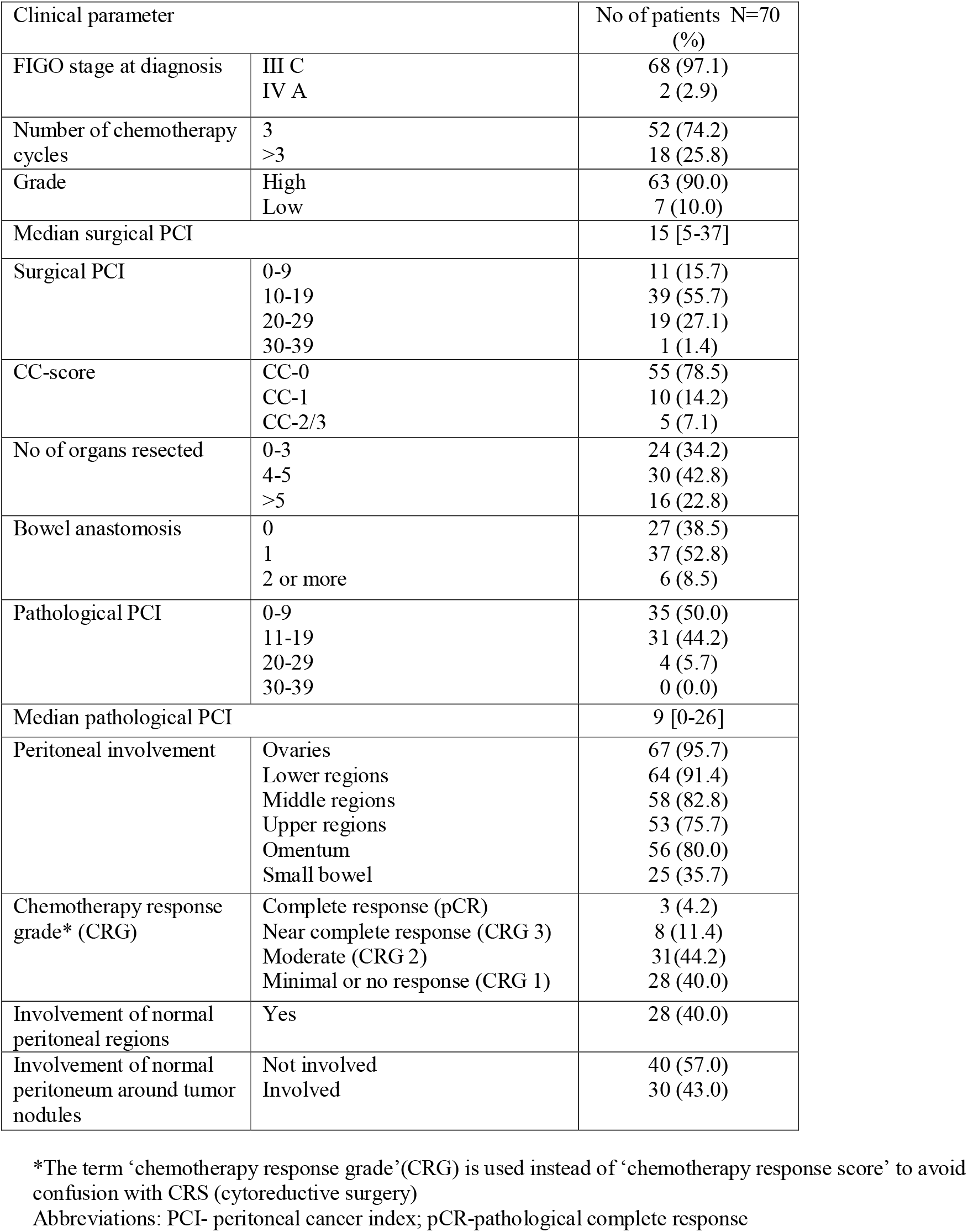
Clinical and surgical findings in 70 patients undergoing interval cytoreductive surgery with total parietal peritonectomy

**Figure 2.**
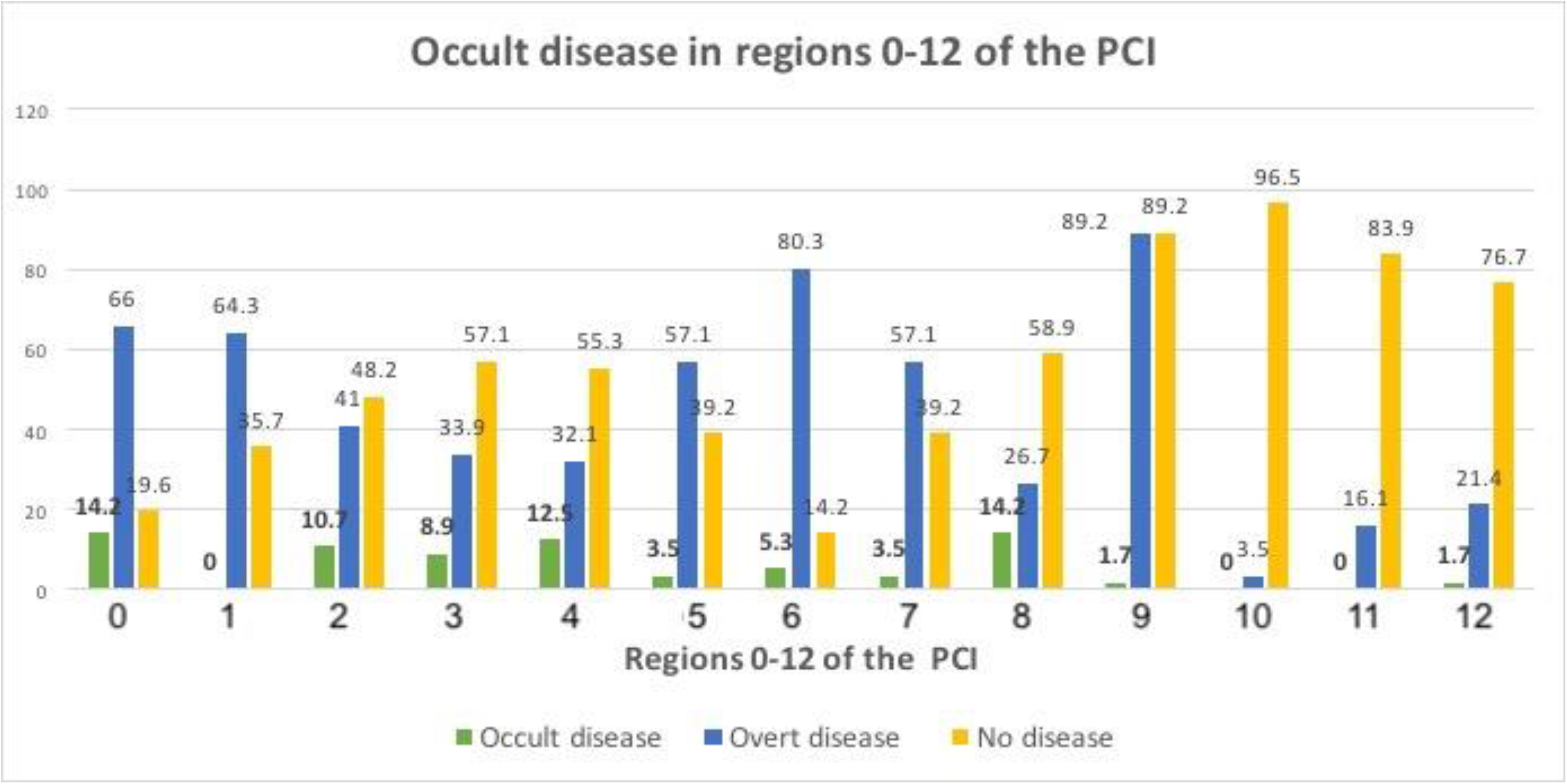
Incidence of occult disease in regions 0-12 of the peritoneal cancer index* *Occult disease is disease in ‘normal appearing’ or involved regions that are given a lesion score of ‘0’ by the surgeon

### Morbidity and mortality

Grade 3-4 complications were seen in 15(21.4%) patients of which 2(2.8%) patients died within 90 days of surgery. One patient died of hemorrhagic shock and another patient died of septic complications. The complications were intraabdominal fluid collections requiring drainage in 6 patients, hemorrhage in 3, intraabdominal sepsis in 2, wound dehiscence in 2 patients, bowel fistula in 1, biliary stricture in 1, urinary tract infection in 1, paralytic ileus in 1, and stoma related complications in 1 patient. Three (4.2%) patients had more than one complication. Grade 1-2 complications were seen in 6 (8.2%) patients. A higher incidence of complications was seen in patients with bowel anastomosis and those receiving post-operative ventilation (**Table 2**).

**Table 2.** Comparison of clinical, surgical and pathological findings in patients with and without complications

| Characteristic |  | All patients<br>N=70 (%) | Patients with<br>complications<br>N=15 (%) | Patients without<br>complications<br>N=55 (%) | p-value |
| --- | --- | --- | --- | --- | --- |
| Number of chemotherapy cycles | 1-3<br>>3 | 52 (74.2)<br>18 (25.8) | 11 (73.3)<br>4 (26.6) | 41 (74.5)<br>14 (25.4) | 0.924 |
| Surgical PCI | 0-9<br>10-19<br>20-29<br>30-39 | 11 (15.7)<br>39 (55.7)<br>19 (27.1)<br>1 (1.4) | 2 (13.3)<br>8 (53.3)<br>4 (26.6)<br>0 (0.0) | 9 (16.3)<br>31 (56.3)<br>15 (27.7)<br>1 (1.8) | 0.785 |
| Bowel anastomosis | 0<br>1<br>>1 | 27 (38.5)<br>37 (52.8)<br>6 (8.5) | 0 (0.0)<br>14 (93.3)<br>1 (6.6) | 27 (49.0)<br>23 (41.8)<br>5 (9.0) | 0.004 |
| Organs resected | 1-3<br>4-5<br>>5 | 24 (34.2)<br>30 (42.8)<br>16 (22.8) | 6 (40.0)<br>6 (40.0)<br>3 (20.0) | 18 (32.7)<br>24 (43.6)<br>13 (23.6) | 0.866 |
| Pathological PCI | 0-9<br>11-19<br>20-29<br>30-39 | 35 (50.0)<br>31 (44.2)<br>4 (5.7)<br>0 (0.0) | 10 (66.6)<br>3 (20.0)<br>2 (13.3)<br>0 (0.0) | 25 (45.4)<br>28 (50.9)<br>2 (3.6)<br>0 (0.0) | 0.062 |
| HIPEC | Yes | 31(44.2) | 7 (46.6) | 24 (43.6) | 0.834 |
| Lymph node dissection | Yes | 54 (77.1) | 14 (93.3) | 40 (72.7) | 0.092 |
| Diaphragm resection | Yes | 9 (12.8) | 0 (0.0) | 9 (16.3) | 0.341 |
| Duration of surgery (minutes) | <300<br>300-540<br>>540 | 14 (20.0)<br>51(72.8)<br>5 (7.1) | 2 (13.3)<br>11 (73.3)<br>2 (13.3) | 12 (21.8)<br>40 (72.7)<br>3 (5.4) | 0.484 |
| Post-operative ventilation | Yes | 41(58.5) | 7 (46.6) | 34 (61.8) | 0.037 |
| Average blood loss (ml) (Mean ± SD) | | 1007±473 | 1025 ±545 | 1003 ± 461 | 0.889 |
| Average duration (mins) (Mean ± SD) | | 393±1170 | 400±146 | 392±112 | 0.847 |
| Average ICU stay (days) (Mean ± SD) | | 3.96±3.35 | 4.77±3.99 | 3.79±3.23 | 0.428 |
| Average hospital stay (days) (Mean ± SD) | | 12.98±5.11 | 13.22±3.49 | 12.93±5.39 | 0.879 |

Abbreviations: PCI-peritoneal cancer index; SD- standard deviation

### Early recurrence and survival

The median follow-up was 13 months [range 0-21 months]. Of 68 evaluable patients, there was no evidence of disease following chemotherapy and in 65 (92.8%) and these were considered to have a complete response. Three patients had persistently high markers and progressive disease despite a CC-1 resection. Early recurrence was seen in 2/65 (3.0%) patients. Thus, 3.0% had platinum resistant disease in the complete responders and 5/70 (7.1%) overall. Overall, 17(24.2%) patients developed recurrence of which 10(14.2%) patients developed recurrence within 12 months of completion of chemotherapy.

The median PFS was 18 months [range 0-21months]

Patients with a lower sPCI and pPCI and with no grade 3-4 complications had a significantly longer PFS (Table 3). A pPCI<15 was the only independent predictor of a longer PFS (p=0.001; hazard ratio 0.126 [95% CI 0.041-0.386]) **(Figure 3)**.

**Table 3.** Cox regression analysis of factors affecting progression-free survival

| Prognostic variable (N) |  | Univariate analysis |  | Multivariate analysis |  |
| --- | --- | --- | --- | --- | --- |
|  |  | Median PFS (months) [95%CI] | p-value | Hazard ratio[95% CI] | p-value |
| Grade | High (63)<br>Low (7) | 18 [14.7-21.2]<br>NR | 0.461 | NA | NA |
| Surgical PCI | <20(50)<br>>20 (20) | NR<br>15 [10.6-19.3] | 0.035 | NA | 0.451 |
| CC-score | CC-0 (55)<br>CC-1-3 (15) | NR<br>17[13.9-20.0] | 0.492 | NA | NA |
| Pathological PCI | <15 (54)<br>>15(16) | NR<br>12 [7.4-16.5] | <0.001 | 0.126<br>(0.041-0.386) | 0.001 |
| HIPEC | Yes (31)<br>No (39) | NR<br>18 [15.1-20.8] | 0.840 | NA | NA |
| Grade 3-4 complications | Yes (15)<br>No (55) | 15[11.6-18.3]<br>NR | 0.056 | NA | 0.203 |
| Lymph node involvement | Yes (22)<br>No (48) | NR<br>18[14.0-21.9] | 0.505 | NA | NA |
| Chemotherapy response grade | 3*(11)<br>1-2 (59) | NR<br>17[13.8-20.1] | 0.570 | NA | NA |
| Disease in 'normal appearing' peritoneum | Yes (28)<br>No (42) | 17<br>18[13.6-22.3] | 0.943 | NA | NA |

Abbreviations: CI-confidence interval; NR-not reached; NA-not applicable
^analysis is limited by a short median follow-up of 13 months
\*Grade 3 includes 3 patients with a pathological complete response

**Figure 3.**
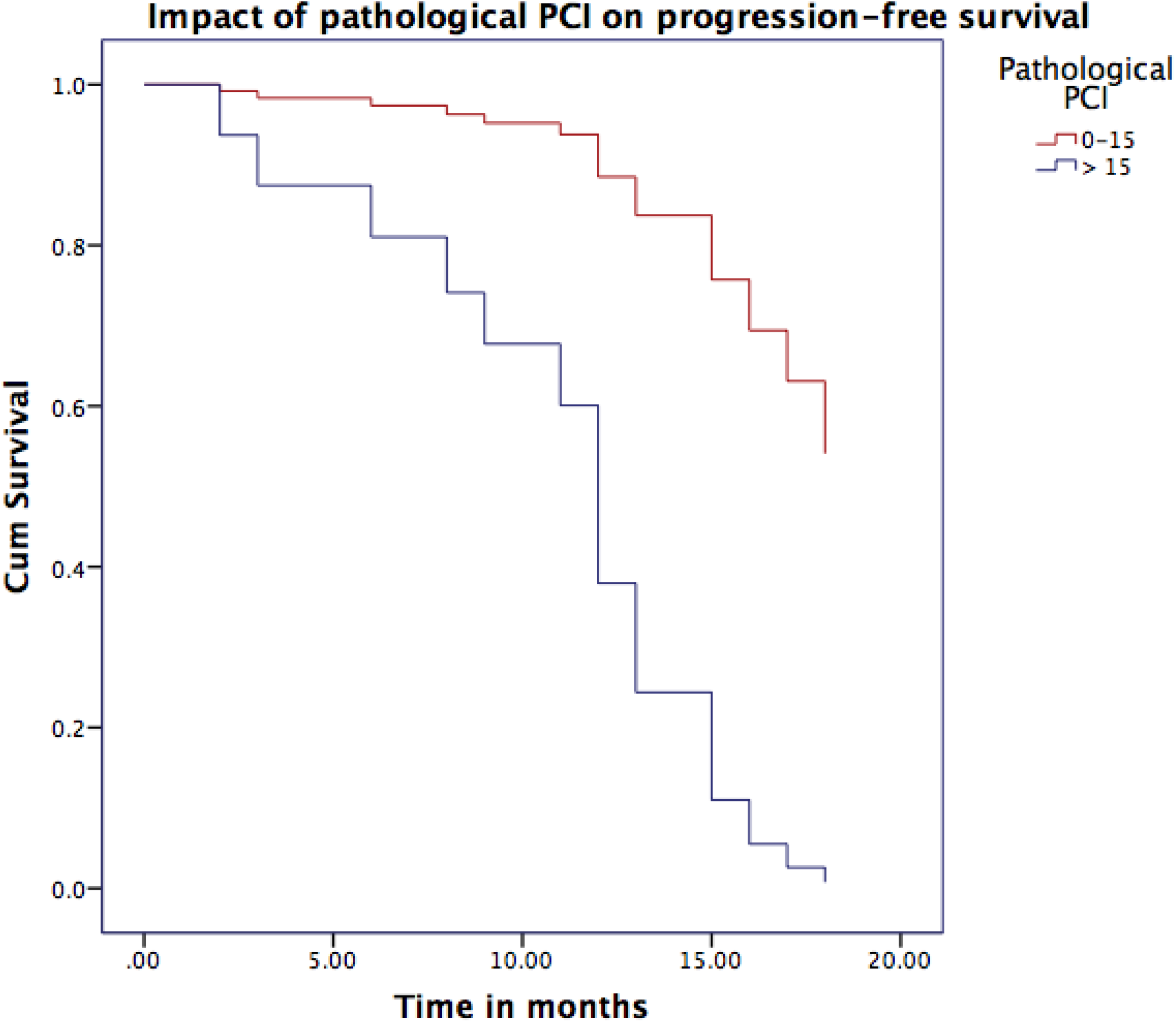
Comparison of progression-free survival in patients with a pathological PCI of more than 15 versus 0-15

## Discussion

This study shows that with more extensive surgery comprising of a TPP, a low incidence of platinum resistance can be achieved in patients undergoing interval CRS. The morbidity and mortality rates can be considered acceptable. The disease extent was quantified using an objective tool, the PCI and patients with a lower PCI experienced a longer PFS. A pathological PCI>15 was the only independent predictor of a shorter PFS. The high incidence of occult disease (40.0%) further supports the rational of performing a TPP (Table 1).

### Platinum resistance

Reported rates of platinum resistance following interval CRS range from 11.3% to 50% with most studies reporting incidences of >30%. [23, 24, 25, 26, 27] Considering only the complete responders, the rate of platinum resistance of 3.0% (7.1% overall) is a significant finding that should be confirmed in a larger series. It demonstrates the importance of addressing occult disease by performing a more extensive surgery targeting sites that were involved before surgery and not just sites of residual disease. Due to the small number of patients, we are not able to study the factors affecting platinum resistance. Even patients with chemotherapy response scores of 1 and 2 did not have a higher incidence of platinum resistance. The reported incidence in these subgroups in around 40%. [21] Considering that over 85% had a chemotherapy response grade of 1/2, early recurrence would be expected in 23 patients. It was observed only in 5(7.1%). The disease extent and major morbidity both had a significant impact on the PFS.

### Residual disease after surgery

Though most investigators agree that no macroscopic residual disease should be the goal, residual disease <1cm is also considered optimal debulking [15, 28]. Residual disease after NACT contains chemotherapy resistant stem cells and should not be considered the same as similar sized residual disease following surgery performed upfront. In this study, 55% had a CC-0 resection. In our practice when there are multiple nodules on the small bowel that are completely resected, the resection is still termed CC-1, not CC-0 since the entire peritoneum has not been removed. Thus, over 90% had a complete gross resection (CC-0/1) which may have been a contributing factor to the low rates of platinum resistance.

### Site of residual disease following NACT

The parietal peritoneum was the commonest site of residual disease which has been shown before too. [29] The small bowel regions were the least common site of residual disease. That combined with the high incidence of occult disease provides further rationale for performing a TPP. The BRCA mutation status of majority of the patients is not known and maintenance therapy was not given to any of the patients. This is the general practice in our country largely due to cost constraints. Though specific data for the NACT group is not available, the incidence of platinum residence was over 35% in the PRIMA trial. [30] Thus, even with maintenance therapy, the incidence of platinum resistance is high. We believe that even in patients who have BRCA mutations and homologous repair deficiency, such extensive surgery will have a benefit that will be additive to what is obtained with maintenance therapies. A larger sample size, a longer follow-up and in future a randomized trial are needed to establish the role of such a procedure in routine practice.

### Multi-organ resection and morbidity

The major morbidity (21.4%) can be considered acceptable. [31, 32, 33] Though 65% had a bowel anastomosis, only one patient who had a bowel fistula. Diverting stoma was performed in <10%. It is our practice to reinforce the rectal anastomosis with a layer of interrupted sutures and avoid performing a diverting stoma as far as possible. [34] The main complication was post-operative fluid collection which could be attributed to lymphadenectomy rather than peritonectomy. These patients underwent fluid aspiration as deemed necessary, usually in the out-patient department. The use of non-invasive ventilation in the post-operative period resulted in no major respiratory complications though all patients had undergone diaphragmatic stripping. The higher morbidity in patients with bowel resection and those receiving post-operative ventilation could be attributed to more extensive disease in them. The two deaths were unexpected. Both hemorrhagic shock and neutropenic sepsis could be attributed to HIPEC. The morbidity should also be evaluated in a larger series.

The relatively small number of patients and short follow-up are the main limitation of this study but the early results are still compelling. This study is prospective in nature, a fixed surgical protocol was followed and all surgeries were performed by surgeons who had performed over 200 CRS procedures.

## Conclusions

Total parietal peritonectomy performed as a part of interval CRS resulted in a very low incidence of platinum resistance in patients with advanced serous ovarian cancer. The post-operative morbidity was acceptable. A pathological PCI>15 was the only independent predictor of a shorter progression-free survival. These findings should be confirmed in a larger series and a randomized trial performed to demonstrate its benefit over the current standard of care that is selective parietal peritonectomy.

## Data Availability

The data will be made available upon reasonable request to the authors

